# Brain-targeted autoimmunity is strongly associated with Long COVID and its chronic fatigue syndrome as well as its affective symptoms

**DOI:** 10.1101/2023.10.04.23296554

**Authors:** Abbas F. Almulla, Michael Maes, Bo Zhou, Hussein K. Al-Hakeim, Aristo Vojdani

**Affiliations:** Department of Psychiatry, Faculty of Medicine, Chulalongkorn University, and King Chulalongkorn Memorial Hospital, the Thai Red Cross Society, Bangkok, Thailand; Medical Laboratory Technology Department, College of Medical Technology, The Islamic University, Najaf, Iraq; Sichuan Provincial Center for Mental Health, Sichuan Provincial People’s Hospital, School of Medicine, University of Electronic Science and Technology of China, Chengdu 610072, China; Key Laboratory of Psychosomatic Medicine, Chinese Academy of Medical Sciences, Chengdu, 610072, China; Cognitive Impairment and Dementia Research Unit, Faculty of Medicine, Chulalongkorn University, Bangkok, Thailand; Department of Psychiatry, Medical University of Plovdiv, Plovdiv, Bulgaria; Research Center, Medical University of Plovdiv, Plovdiv, Bulgaria; Kyung Hee University, 26 Kyungheedae-ro, Dongdaemun-gu, Seoul 02447, Korea; Department of Chemistry, College of Science, University of Kufa, Kufa, Iraq; Immunosciences Lab, Inc., Los Angeles, CA 90035, USA; Cyrex Laboratories, LLC, Phoenix, AZ 85034, USA

**Keywords:** Long COVID, neuroimmune, chronic fatigue syndrome, depression, affective disorders, oxidative stress

## Abstract

**Background:** Autoimmune responses contribute to the pathophysiology of Long COVID, affective symptoms and myalgic encephalomyelitis/chronic fatigue syndrome (ME/CFS).

**Objectives:** To examine whether Long COVID, and its accompanying affective symptoms and CFS are associated with immunoglobulin (Ig)A/IgM/IgG directed at neuronal proteins including myelin basic protein (MBP), myelin oligodendrocyte glycoprotein (MOG), synapsin, α+β-tubulin, neurofilament protein (NFP), cerebellar protein-2 (CP2), and the blood-brain-barrier-brain-damage (BBD) proteins claudin-5 and S100B.

**Methods:** IgA**/**IgM/IgG to the above neuronal proteins, human herpes virus-6 (HHV-6) and Severe Acute Respiratory Syndrome Coronavirus 2 (SARS-CoV-2) were measured in 90 Long COVID patients and 90 healthy controls, while C-reactive protein (CRP), and advanced oxidation protein products (AOPP) in association with affective and CFS ratings were additionally assessed in a subgroup thereof.

**Results:** Long COVID is associated with significant increases in IgG directed at tubulin (IgG-tubulin), MBP, MOG and synapsin; IgM-MBP, MOG, CP2, synapsin and BBD; and IgA-CP2 and synapsin. IgM-SARS-CoV-2 and IgM-HHV-6 antibody titers were significantly correlated with IgA/IgG/IgM-tubulin and -CP2, IgG/IgM-BBD, IgM-MOG, IgA/IgM-NFP, and IgG/IgM-synapsin. Binary logistic regression analysis shows that IgM-MBP and IgG-MBP are the best predictors of Long COVID. Multiple regression analysis shows that IgG-MOG, CRP and AOPP explain together 41.7% of the variance in the severity of CFS. Neural network analysis shows that IgM-synapsin, IgA-MBP, IgG-MOG, IgA-synapsin, IgA-CP2, IgG-MBP and CRP are the most important predictors of affective symptoms due to Long COVID with a predictive accuracy of r=0.801.

**Conclusion:** Brain-targeted autoimmunity contributes significantly to the pathogenesis of Long COVID and the severity of its physio-affective phenome.

## Introduction

Ongoing neuro-psychiatric symptoms have been reported in a substantial percentage of coronavirus disease (COVID) survivors (Davis, McCorkell et al. 2023). These symptoms encompass chronic fatigue syndrome (CFS), depression, and anxiety persisting for up to 12 months post-recovery, commonly known as “Long COVID disease” (Groff, Sun et al. 2021, Lopez-Leon, Wegman-Ostrosky et al. 2021, Premraj, Kannapadi et al. 2022). Recent data reveal that globally, at least 65 million individuals suffer from Long COVID disease (Davis, McCorkell et al. 2023).

In a recent previous article co-authored by some of the present’s study collaborators, it was observed that CFS and affective symptoms due to Long COVID are predicted by elevated peak body temperature (PBT) and decreased oxygen saturation (SpO2) during the acute phase of illness (Al-Hadrawi, Al-Rubaye et al. 2022). Both PBT and lower SpO2 are indices of the severity of the immune-inflammatory response during acute infection (Al-Jassas, Al-Hakeim et al. 2022). In another article by some of this study’s authors, a validated latent vector could be extracted from the CFS, fibromyalgia, depressive and anxiety symptoms due to acute and Long COVID and this latent construct was named the “physio-affective phenome” of acute COVID-19 (Al-Jassas, Al-Hakeim et al. 2022) or Long COVID (Al-Hadrawi, Al-Rubaye et al. 2022, Al-Hakeim, Al-Rubaye et al. 2022, Al-Hakeim, Al-Rubaye et al. 2022, Maes, Al-Rubaye et al. 2022).

However, there is still much debate on what causes Long COVID disease and the severity of CFS, depression and anxiety symptoms due to Long COVID. In this context, in yet another study, some of the same authors have identified molecular pathways implicated in the onset of symptoms in individuals with Long COVID disease, including activation of immune-inflammatory processes with oxidative and nitrosative stress reactions (Al-Hakeim, Al-Rubaye et al. 2022, Al-Hakeim, Al-Rubaye et al. 2022), increased insulin resistance (Al-Hakeim, Al-Rubaye et al. 2023, Maes, Almulla et al. 2023), decreased tryptophan levels and increased tryptophan catabolites, such as kynurenine (Al-Hakeim, Abed et al. 2023, Al-Hakeim, Khairi Abed et al. 2023). Moreover, a recent meta-analysis reported that Long COVID disease is accompanied by increased C-reactive protein (CRP), D-dimer, lactate dehydrogenase, leukocytes, lymphocytes, and interleukin (IL)-6 (Yong, Halim et al. 2023).

Recently, Vojdani et al. discovered that Long COVID patients show elevated levels of immunoglobulins (Ig) IgG/IgM directed against Severe Acute Respiratory Syndrome Coronavirus 2 (IgG/IgM-SARS-CoV-2), human Herpesvirus type 6 (HHV-6) and its deoxyuridine 5′-triphosphate nucleotidohydrolase (HHV-6-duTPase) along with IgA/IgM at activin-A (a self-antigen) (Vojdani, Almulla et al. 2023). These findings confirmed previous studies which indicated that Long COVID disease is accompanied by persistence of SARS-CoV-2, reactivation of dormant viruses, and autoimmune reactions against self-proteins (Acosta-Ampudia, Monsalve et al. 2022, Rojas, Rodríguez et al. 2022, Su, Yuan et al. 2022, Vojdani, Vojdani et al. 2023). Importantly, Vojdani et al. were able to predict the Long COVID diagnosis with high sensitivity (78.9%) and specificity (81.8%) based on elevated levels of IgA-activin-A, IgG-HHV-6, IgM-HHV-6-duTPase, IgG-SARS-CoV-2, and IgM-HHV-6, and a factor extracted from all IgA levels to all viral antigens (Vojdani, Almulla et al. 2023).

Neurological disease, which involves both the central and peripheral nervous systems, is observed in more than one-third of patients with COVID-19 and long COVID (Stefanou, Palaiodimou et al. 2022). The entry of SARS-CoV-2 into central nervous system (CNS) cells is facilitated via the engagement of the virus with the angiotensin-converting enzyme (ACE) receptor on the surface of neurons, endothelial and smooth muscle cells of the cerebral blood vessels (Stefanou, Palaiodimou et al. 2022). This entry of the virus, and the entry of T helper (Th)-1 and Th-17 cells, as well M1 macrophage cytokines into the CNS may result in activation of microglia, resulting in neuronal cell damage, the release of neuronal cell antigens, and antibody production (Elizalde-Díaz, Miranda-Narváez et al. 2022, Stefanou, Palaiodimou et al. 2022).

Autoantibodies that are directed to endogenous proteins have been observed in several neuro-psychiatric illnesses which show clinical and pathophysiological features like Long COVID (Morris and Maes 2013, Apostolou, Rizwan et al. 2022, Komaroff and Lipkin 2023). For example, antibodies against synapsin in psychiatric patients are detected in association with increased agitation (Sæther, Vaaler et al. 2019). Increased neurofilament light and P-tau concentrations were established in patients with depression (Al-Hakeim, Al-Naqeeb et al. 2023). Increased levels of IgG directed at myelin basic protein (MBP, essential for the myelin sheath in brain oligodendrocytes) have been identified in bipolar patients (Kamaeva, Smirnova et al. 2022). CFS patients display increased antibodies against a multitude of neuronal proteins including microtubulin-related protein-2, a component of the cytoskeleton in eukaryotic cells (Vernon and Reeves 2005, Morris and Maes 2013).

Numerous studies reported that individuals recovered from COVID-19 (3-12 months after infection) show significant elevations of specific autoantibodies including those against calprotectin, nucleoprotein, whole spike, and spike subunits (Moody, Sonda et al. 2022), ACE2 (Arthur, Forrest et al. 2021), and apolipoprotein A-1 (Apo-A1) (L’Huillier, Pagano et al. 2022). Additionally IgG/IgM against cardiolipin and betaL2 glycoprotein I (Pisareva, Badiou et al. 2023), cyclic citrullinated peptide (CCP) and tissue transglutaminase (Lingel, Meltendorf et al. 2021) along with IgG against interferon (IgG-IFN), histone and centromere protein were also detected (Rojas, Rodríguez et al. 2022). There is also evidence that the blood-brain-barrier (BBB) is dysfunctional in Long COVID (Krasemann, Haferkamp et al. 2022, Hernández-Parra, Reyes-Hernández et al. 2023). Increased autoantibody titers to BBB proteins such as claudin are observed in Long COVID patients (Fonseca, Filgueiras et al. 2023). In the latter, the severity of illness is associated with increased serum levels of S100B, a danger-associated molecular pattern (DAMP) molecule, suggesting increased BBB permeability and brain damage (Aceti, Margarucci et al. 2020).

However, it is largely unknown whether autoimmune reactions directed against neuronal proteins are a feature of Long COVID disease. Hence, the aim of this study is to examine autoantibodies (IgA/IgM/IgG) directed at MBP, myelin oligodendrocyte glycoprotein (MOG), cerebellar-protein-2, synapsin, tubulin, neurofilament protein (NFP), and BBB-brain damage (BBD) proteins (claudin-5 and S100B) in Long COVID disease. In addition, we employ the precision medicine method (Maes 2022, Maes and Stoyanov 2022) to delineate whether these antibodies can predict CFS and affective symptoms due to Long COVID disease.

## Participants and Methods

### Participants

The World Health Organization (WHO) criteria for Long COVID disease (World Health Organization 2021) were followed by experienced clinicians when recruiting patients in the present study. Based on these criteria, patients with long COVID are those who suffered from a verified COVID-19 infection and experienced at least two of the following symptoms for at least two months after the initial infection: fatigue, memory or concentration difficulties, muscle discomfort, loss of olfactory or gustatory senses, emotional distress, and cognitive dysfunction. These symptoms may persist beyond the initial acute phase of the illness or become apparent 2-3 months following the initial infection. Subsequently, we recruited 90 patients with Long COVID and 90 healthy controls. Two distinct research methodologies and study samples were applied in the study. The first part (Part 1) is a retrospective case-control study design encompassing 58 Long COVID patients and 14 normal controls to delineate the impact of PBT and SpO2 during the acute phase of illness on the autoimmune biomarkers and the associations between the latter and neuropsychiatric rating scales. In addition, the duration of the acute infectious phase on the biomarkers and their effect on the symptoms of Long COVID were examined. A second case-control study (Part 2) with a sample size of 180 (90 Long COVID and 90 controls) was conducted to investigate the association between biomarkers and diagnosis (long COVID versus healthy controls).

Part 1 of the study included Iraqi participants whose acute COVID-19 stage was diagnosed by professional virologists and clinicians. Those participants sought medical care from various healthcare facilities, including Imam Sajjad Hospital, Hassan Halos Al-Hatmy Hospital for Infectious Diseases, Middle Euphrates Oncology Center, Al-Najaf Educational Hospital, and Al-Sader Medical City, all located in Najaf, Iraq. The Long COVID patients included here showed persistent characteristics of Long COVID symptoms extending for 12-16 weeks or more and suffered before from acute infection as diagnosed using the presence of fever, cough, respiratory distress, and anosmia or ageusia, an affirmative reverse transcription real-time polymerase chain reaction (rRT-PCR) test, and the detection of IgM antibodies against SARS-CoV-2 during the initial phase of the disease. Control participation was not accepted if controls showed a) indications reminiscent of COVID-19 or any other infectious ailment, b) a confirmatory rRT-PCR test result, or heightened levels of IgM antibodies against SARS-CoV-2. Moreover, we excluded subjects who reported a history of major depressive episodes, bipolar disorder, dysthymia, generalized anxiety disorder, panic disorder, schizo-affective disorder, schizophrenia, psycho-organic syndromes, or substance use disorders (except nicotine dependence). Additionally, patients and controls with neuro-immune, autoimmune, and immune-related conditions, encompassing Parkinson’s disease, CFS, Alzheimer’s disease, multiple sclerosis, stroke, psoriasis, chronic kidney disease, Chronic obstructive pulmonary disease (COPD), and scleroderma were not eligible to participate. Lactating or pregnant women were also not considered for inclusion.

Fifty-four COVID patients who were investigated in Cyrex Laboratory in California were recruited for part 2 of this study. Those patients were primarily admitted with symptoms of fatigue, cognitive impairment, neurocognitive deterioration, dyspnea, headache or vertigo, sleep disturbances, persistent cough, thoracic discomfort, musculoskeletal pain, gastrointestinal complications, and menstrual irregularities that minimally lasted for 3 months beyond their acute COVID-19 stage. Everyone was tested for elevated levels of IgG antibodies against the spike protein and nucleoprotein of SARS-CoV-2. Furthermore, Innovative Research, located in Novi, Michigan, provided 76 control sera samples from pre-COVID individuals free of known health conditions. Additionally, it has been confirmed that they weren’t infected with human immunodeficiency virus (HIV) and hepatitis C. SARS-CoV-2 IgG antibody obtained from Zeus Scientific was employed to verify that all control samples show negative tests. All serum samples were preserved at −20°C pending their utilization in the antibody tests.

The Ethics Committee of the College of Medical Technology at the Islamic University of Najaf in Iraq released the approval to carry out this research (Document No: 34/2023). Before their inclusion in the research, all participants or their legal representatives provided written informed consent. The study’s design and implementation adhered to the International Conference on Harmonization of Good Clinical Practice standards, the Belmont Report, the Council of International Organizations of Medicine (CIOMS) Guideline, and Iraqi and global ethical and privacy regulations. The institutional review board of our institution aligns with the International Guidelines for the Conduct of Safe Human Research (ICH-GCP).

### Clinical measurements

A professional psychiatrist interviewed all participants in study 1 around 3-4 months after recovery from the acute phase of SARS-CoV-2 infection. Data, including socio-demographic and clinical features, were obtained in this interview from individuals who participated in this study. The same psychiatrist employed the Fibro-Fatigue scale to assess the severity of CSF (Zachrisson, Regland et al. 2002). The severity of depressive symptoms was evaluated utilizing two established measures, namely the Hamilton Depression Rating Scale (Hamilton 1960) and the Beck Depression Inventory-II (BDI) (Hautzinger 2009). The Hamilton Anxiety Rating Scale (HAMA) (Hamilton 1959) was employed to examine severity of anxiety symptoms. The severity of the Long COVID phenome was ascertained by deriving a z unit-based composite score, formulated as z HAMD + z HAMA + z BDI, denoted as Comp_affective. Diagnostic criteria for Tobacco Use Disorder (TUD) were obtained from the Diagnostic and Statistical Manual of Mental Disorders, Fifth Edition (DSM-5). The body mass index (BMI) was calculated by taking an individual’s weight (expressed in kilograms) and dividing it by the square of their height (measured in meters). Medical records were examined by clinicians to retrieve SpO2, PBT, and duration of illness of patients. Using a digital sublingual thermometer with an audible signal and an electronic oximeter, both manufactured by Shenzhen Jumper Medical Equipment Co. Ltd., a skilled paramedical specialist recorded these readings. The was estimated from their medical records.

### Assays

Early in the morning (7:30-9:00 a.m.), five mL of venous blood samples were taken from fasting participants, which were then transferred into clear, sterile serum tubes while omitting any hemolyzed sample. Following a ten-minute clotting period, the samples underwent centrifugation for five minutes at a rotational speed of 3000 revolutions per minute (rpm). Subsequently, the resultant serum was meticulously transferred into multiple new Eppendorf tubes. The CRP latex slide test, a product manufactured by Spinreact® in Barcelona, Spain, was utilized to conduct measurements of CRP in human serum. The advanced oxidation protein products (AOPP) in serum were quantified using enzyme-linked immunosorbent assay (ELISA) kits from Nanjing Pars Biochem Co., Ltd. in Nanjing, China. The computation of the Homeostatic Model Assessment for Insulin Resistance (HOMA2-IR) was carried out by HOMA2 calculator accessible at https://www.dtu.ox.ac.uk/homacalculator, which serves as a metric for evaluating insulin resistance, involved the utilization of fasting insulin and serum glucose levels.

### Antigens

MBP was purchased from Sigma-Aldrich (St. Louis, MO, USA), while Bio-Synthesis (Lewisville, TX, USA) synthesized the MOG peptide. Additionally, NFP and synapsin were obtained from Bio-Techne R & D Systems (Minneapolis, MN, USA) and Antibodies (Limerick, PA, USA), respectively. Cerebellar protein-2 (CP2) was purchased from CUSABIO (Houston, TX, USA) and α and β tubulin from Abcam (Cambridge, MA, USA). The two major BBD proteins, claudin-5 and S100B, were synthesized by Bio-Synthesis® (Lewisville, TX, USA).

### Antibody measurements

ELISA was employed to detect serum antibodies directed against neuronal antigens, including tubulin, BBD, cerebellar-protein-2, MBP, MOG, NFP, and synapsin. A multiple step procedure was used: a) proteins and peptides were solubilized using a Tris buffer with a pH of 7.2. Subsequently, 100 ml of each solution was allocated to microwell plates in concentrations varying between 0.5 to 1 microgram, using a 0.1 M carbonate buffer with a pH of 9.6; b) these plates were subjected to incubation at ambient temperature (25°C) for 16 hours, followed by a cooling period of 8 hours; c) post removal of the plate contents, a triple wash was conducted using a 250 ml solution consisting of (0.01 M PBS with a pH of 7.4 and 0.05% Tween 20); d) to counteract the non-specific adherence of serum immunoglobulins, the plates were re-incubated post the addition of 250 ml of a solution, which comprised 2% bovine serum albumin (BSA) and 2% dry milk in the PBS buffer to each well; e) following washing the microtiter plates four times with PBS buffer, we added serum dilutions of 1:100 for IgG and IgM antibodies and 1:50 for IgA antibodies to duplicate wells; f) after a one-hour incubation at 25°C, the ELISA plates underwent a quintuple wash with PBS buffer. Alkaline phosphatase-labeled anti-human IgG (diluted 1:800), anti-human IgM (diluted 1:600), and anti-human IgA (diluted 1:200) antibodies were then diluted and 100 ml of each were introduced to the pertinent plate sets; g) following additional washing, 100 ml of the substrate was added, and the color development was arrested using 50 ml of 1 N NaOH. A designated ELISA reader, calibrated to 405 nm, read the color intensity. Sera from Long COVID patients with recognized antibody titers were employed as positive controls. At the same time, wells coated with BSA, human serum albumin (HAS), and fetal bovine serum functioned as negative controls or blanks. The ELISA optical density (OD)s was translated to indices after the deduction of the background OD from both the sample and calibrator ODs, utilizing a specific formula outlined below:

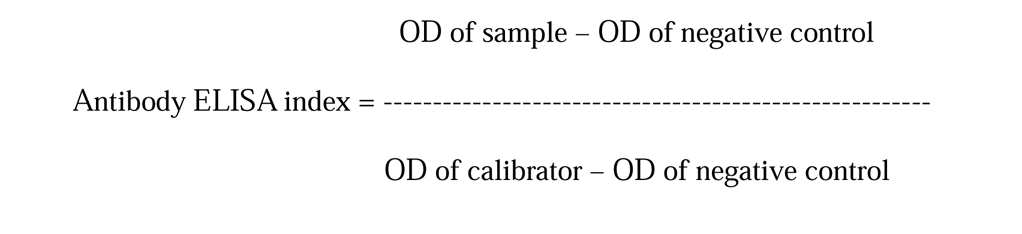

As described by Vojdani et al. (Vojdani, Almulla et al. 2023) we assayed IgA/IgM/IgG levels to the SARS-CoV-2, HHV-6, HHV-6-duTPAse, Epstein-Barr Virus (EBV), and activin-A. In the current study, we used the most important predictors, which were established by Vojdani et al. (2023), in order to improve the prediction of Long COVID and its phenome based on the neuronal biomarkers measured in the current study, namely IgG-SARS-CoV-2, IgG-HHV6, IgA-Activin A and a principal component extracted from IgA directed to SARS-CoV-2, HHV-6, HHV6-duTPase, EBV, and EBV-duTPase. The latter index of IgA protection against viral infections is significantly decreased in Long COVID (Vojdani, Almulla et al. 2023).

### Statistical analysis

In the current study, we conducted an analysis of variance (ANOVA) to compare continuous variables among study groups, while a contingency table analysis compared category variables. Pearson’s correlation coefficients were used to analyze the relationships between IgA/IgM/IgG and neuro-psychiatric symptoms scales, PBT and AOPP. Our study employed binary logistic regression analysis to assess the relationship between IgA/IgG/IgM responses and Long COVID diagnosis, using healthy controls as the reference category. In these analyses, we have adjusted for potential confounders such as age, sex, and study location. We computed B (standard error, SE), Wald statistics with p-values, the Odds ratio with 95% confidence intervals (CI), the classification table, and Nagelkerke pseudo-R square (used for estimating the effect size). The continuous data from the rating scale scores were predicted using neural networks with the biomarkers as independent variables. To determine the primary predictors of CFS and affective symptoms in Long COVID, we conducted multivariate regression analyses while adjusting for age, gender, and BMI. A stepwise automated approach was employed, with p-values of 0.05 and 0.10 serving as entry and exclusion criteria, respectively. Key model metrics, such as F, df, and p-values, along with the total variance (R^2^) and standardized beta coefficients, were calculated. We evaluated the variance inflation factor (VIF) and tolerance to address potential collinearity issues. To evaluate heteroskedasticity, we utilized the White and modified Breusch-Pagan tests. All tests adopted a significance threshold set at a p-value of 0.05 and were conducted in a two-tailed manner.

Our multilayer perceptron neural network models used the most important IgG, IgM, and IgA responses to the antigens, together with age, gender, education level, AOPP, HOMA2-IR, and CRP values as input variables. Up to eight nodes were used in an automated feedforward network model with one or two hidden layers. The models were trained using a maximum of 250 epochs in a batch-type training session. The termination criterion was established based on a consecutive step failing to reduce the error term further. The error, relative error, and the proportion of misclassifications were computed, or we computed the models’ predictive precision through the coefficient of determination (R^2^) comparing predicted versus observed values. The significance and relative prominence of the input variables were evaluated and represented in an importance chart.

For feature reduction purposes, Principal Component Analysis (PCA) was applied. A Principal Component (PC) was deemed validated when the explained variance (EV) reached or exceeded 50%, the anti-image correlation matrix was satisfactory, factorability indicators were appropriate with a Kaiser-Meyer-Olkin (KMO) value surpassing 0.65, Bartlett’s test of sphericity was significant, and all PC loadings exceeded 0.7. The latest Windows version of IBM’s SPSS program (SPSS 29) was used for all statistical tests.

An a priori power analysis, conducted using G*Power 3.1.9.7, indicates that the required sample size should be no less than 126 subjects to discern variations in a chi-square test, assuming an effect size of 0.25, a significance level (p) of 0.05, a power of 0.8, and degrees of freedom (df) equal to 1.

## Results

### Socio-demographic and clinical characteristics of Long COVID

**Table 1** in our study delineates the socio-demographic details, PBT, SpO2, and the duration of the acute COVID phase in patients with Long COVID versus controls. Additionally, it presents scores from diverse psychiatric evaluations and biomarker assessments, encompassing FF, HAMA, HAMD, BDI, CRP, AOPP, and HOMA2-IR index. Except for residence, there were no statistically significant differences in the social-demographic features across the research groups. In their acute infection stage, patients exhibited a significant increase in PBT and a marked decrease in SpO2 compared to the control group. In the Long COVID cohort, the mean duration of the acute infectious phase was 14.1 days (standard deviation, 5.6 days). Individuals diagnosed with Long COVID exhibited significantly elevated mean scores on the FF, HAMA, BDI, and HAMD scales. Additionally, those patients demonstrated increased CRP, AOPP, and HOMA2IR concentrations compared to the healthy control group.

**Table 1:**
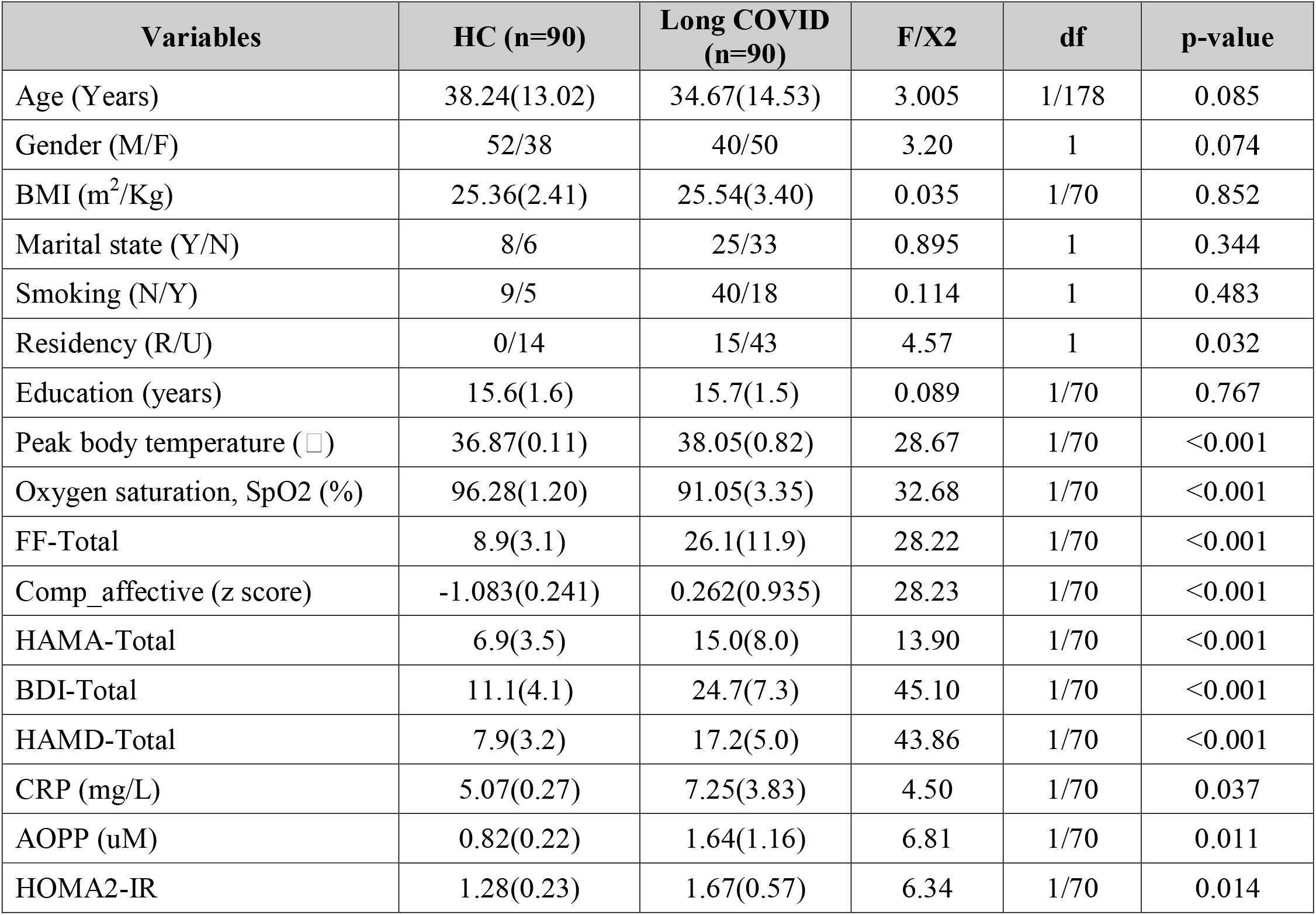
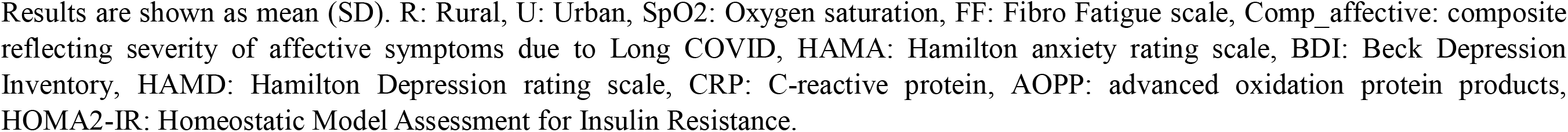
Socio-demographic and clinical data, body temperature, oxygen saturation (SpO2), and neuro-psychiatric rating scales in healthy controls (HC) and Long COVID patients.

| Variables | HC (n=90) | Long COVID (n=90) | F/X<sup>2</sup> | df | p-value |
| --- | --- | --- | --- | --- | --- |
| Age (Years) | 38.24(13.02) | 34.67(14.53) | 3.005 | 1/178 | 0.085 |
| Gender (M/F) | 52/38 | 40/50 | 3.20 | 1 | 0.074 |
| BMI (m<sup>2</sup>/Kg) | 25.36(2.41) | 25.54(3.40) | 0.035 | 1/70 | 0.852 |
| Marital state (Y/N) | 8/6 | 25/33 | 0.895 | 1 | 0.344 |
| Smoking (N/Y) | 9/5 | 40/18 | 0.114 | 1 | 0.483 |
| Residency (R/U) | 0/14 | 15/43 | 4.57 | 1 | 0.032 |
| Education (years) | 15.6(1.6) | 15.7(1.5) | 0.089 | 1/70 | 0.767 |
| Peak body temperature (°C) | 36.87(0.11) | 38.05(0.82) | 28.67 | 1/70 | <0.001 |
| Oxygen saturation, SpO<sub>2</sub> (%) | 96.28(1.20) | 91.05(3.35) | 32.68 | 1/70 | <0.001 |
| FF-Total | 8.9(3.1) | 26.1(11.9) | 28.22 | 1/70 | <0.001 |
| Comp_affective (z score) | -1.083(0.241) | 0.262(0.935) | 28.23 | 1/70 | <0.001 |
| HAMA-Total | 6.9(3.5) | 15.0(8.0) | 13.90 | 1/70 | <0.001 |
| BDI-Total | 11.1(4.1) | 24.7(7.3) | 45.10 | 1/70 | <0.001 |
| HAMD-Total | 7.9(3.2) | 17.2(5.0) | 43.86 | 1/70 | <0.001 |
| CRP (mg/L) | 5.07(0.27) | 7.25(3.83) | 4.50 | 1/70 | 0.037 |
| AOPP (uM) | 0.82(0.22) | 1.64(1.16) | 6.81 | 1/70 | 0.011 |
| HOMA2-IR | 1.28(0.23) | 1.67(0.57) | 6.34 | 1/70 | 0.014 |

Results are shown as mean (SD). R: Rural, U: Urban, SpO<sub>2</sub>: Oxygen saturation, FF: Fibro Fatigue scale, Comp\_affective: composite reflecting severity of affective symptoms due to Long COVID, HAMA: Hamilton anxiety rating scale, BDI: Beck Depression Inventory, HAMD: Hamilton Depression rating scale, CRP: C-reactive protein, AOPP: advanced oxidation protein products, HOMA2-IR: Homeostatic Model Assessment for Insulin Resistance.

### Association between Long COVID disease and autoimmune mediators

We utilized binary logistic regression analysis to determine the most prominent variables in predicting the occurrence of Long COVID disease. Our study’s dependent variable was Long COVID diagnosis, with the control group as the reference group. The independent variables included in this study were the IgA, IgG, and IgM responses to the neuronal antigens. Statistical adjustments were performed to account for age, sex, and research site, as outlined in **Table 2**. Regression #1 shows that IgG-Tubulin is significantly and positively associated with Long COVID disease with an effect size of 0.064. IgM-BBD (regression #2) has a significant positive association with Long COVID disease with an effect size of 0.085. Regression #3 and #4 reveal that Long COVID showed significant and positive associations with IgA and IgM against CP2, and the effect sizes were 0.049 and 0.028, respectively. Long COVID disease is significantly and positively associated with IgG and IgM against MBP, as shown in regression #5 and #6 with effect sizes of 0.152 and 0.216, respectively. Regressions #7 and #8 show that IgG and IgM (with effect sizes 0.143 and 0.061, respectively) directed at MOG have positive significant associations with Long COVID disease. Our results (regression #9, #10 and #11) indicate that IgA, IgM, and IgG at synapsin have positive and significant associations with Long COVID disease, with effect sizes of 0.073, 0.080 and 0.049, respectively. **Figure 1** shows the significant increases in IgA, IgM, and IgG towards synapsin in patients with Long COVID versus healthy controls. We have computed the sum of all IgA and IgG OD values to neuronal antigens and used these as an overall index of brain reactive autoantibodies (IgA-BRA and IgG-BRA). IgG-BRA in regression #12 was significantly and positively associated with Long COVID disease, with 0.080 as the effect size.

**Figure 1.**
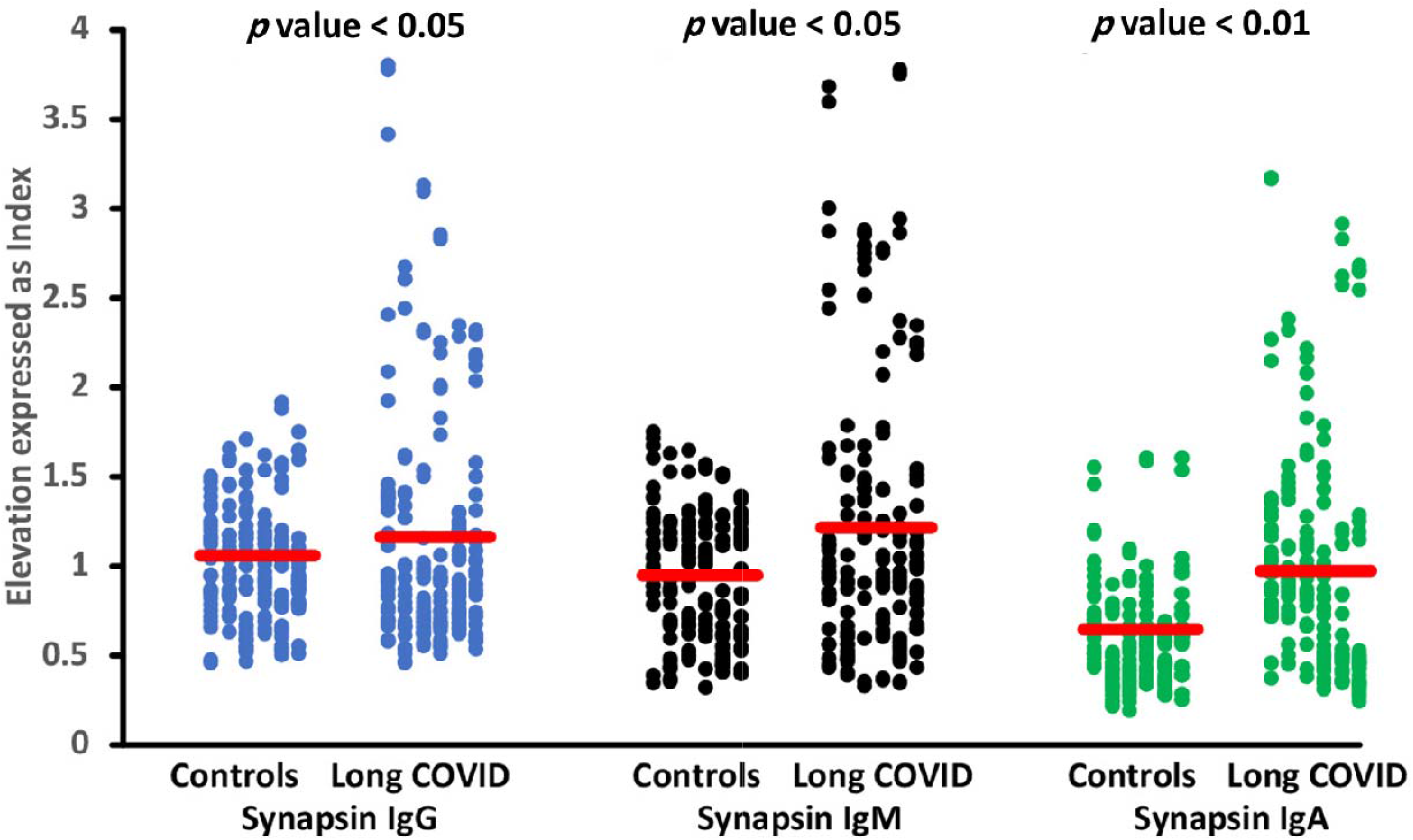
Differences in immunoglobulins IgM, IgA and IgG levels directed against synapsin between ID disease patients and healthy controls.

**Table 2.**
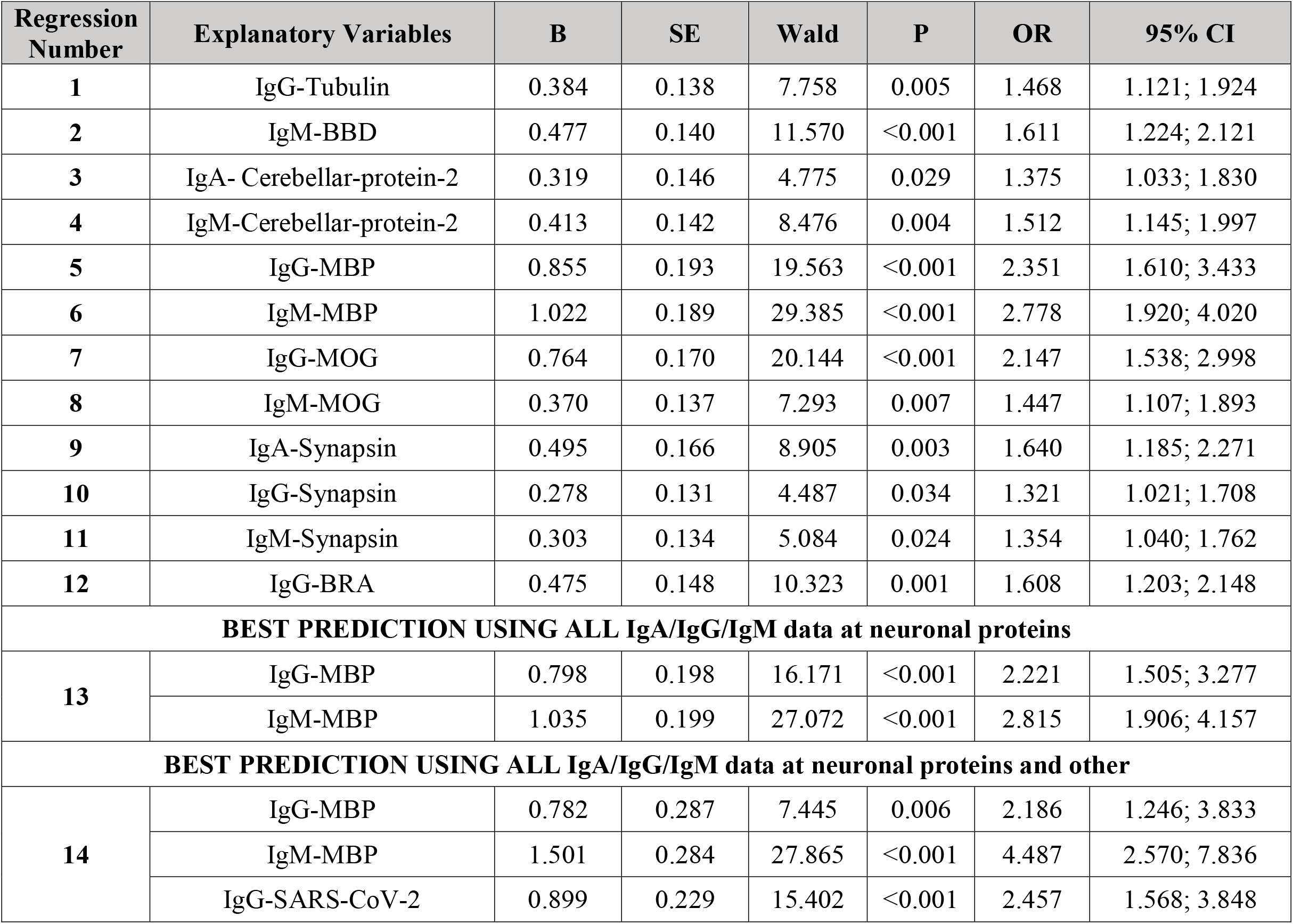
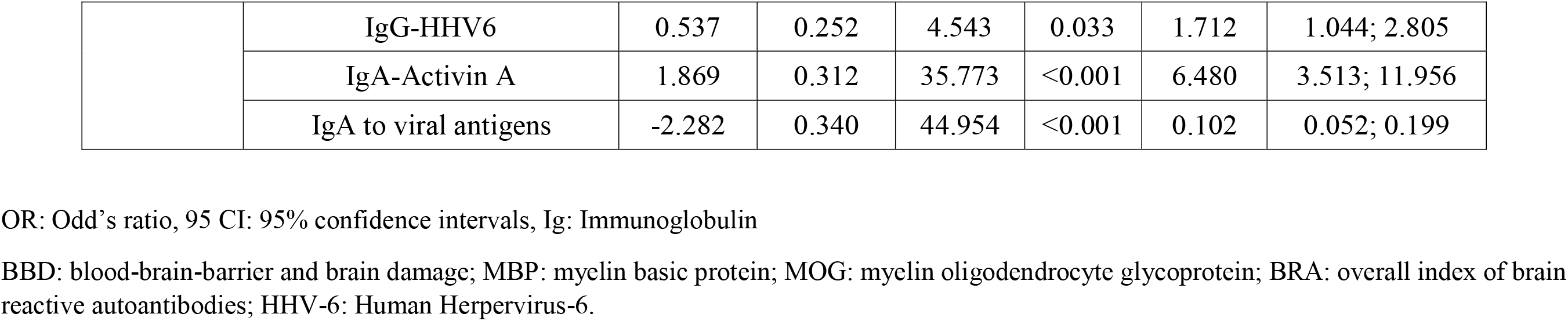
Results of binary logistic regression analysis with the diagnosis Long COVID as dependent variable (heathy controls as reference group).

| Regression Number | Explanatory Variables | B | SE | Wald | P | OR | 95% CI |
| --- | --- | --- | --- | --- | --- | --- | --- |
| 1 | IgG-Tubulin | 0.384 | 0.138 | 7.758 | 0.005 | 1.468 | 1.121; 1.924 |
| 2 | IgM-BBD | 0.477 | 0.140 | 11.570 | <0.001 | 1.611 | 1.224; 2.121 |
| 3 | IgA- Cerebellar-protein-2 | 0.319 | 0.146 | 4.775 | 0.029 | 1.375 | 1.033; 1.830 |
| 4 | IgM-Cerebellar-protein-2 | 0.413 | 0.142 | 8.476 | 0.004 | 1.512 | 1.145; 1.997 |
| 5 | IgG-MBP | 0.855 | 0.193 | 19.563 | <0.001 | 2.351 | 1.610; 3.433 |
| 6 | IgM-MBP | 1.022 | 0.189 | 29.385 | <0.001 | 2.778 | 1.920; 4.020 |
| 7 | IgG-MOG | 0.764 | 0.170 | 20.144 | <0.001 | 2.147 | 1.538; 2.998 |
| 8 | IgM-MOG | 0.370 | 0.137 | 7.293 | 0.007 | 1.447 | 1.107; 1.893 |
| 9 | IgA-Synapsin | 0.495 | 0.166 | 8.905 | 0.003 | 1.640 | 1.185; 2.271 |
| 10 | IgG-Synapsin | 0.278 | 0.131 | 4.487 | 0.034 | 1.321 | 1.021; 1.708 |
| 11 | IgM-Synapsin | 0.303 | 0.134 | 5.084 | 0.024 | 1.354 | 1.040; 1.762 |
| 12 | IgG-BRA | 0.475 | 0.148 | 10.323 | 0.001 | 1.608 | 1.203; 2.148 |
| **BEST PREDICTION USING ALL IgA/IgG/IgM data at neuronal proteins** |  |  |  |  |  |  |  |
| 13 | IgG-MBP | 0.798 | 0.198 | 16.171 | <0.001 | 2.221 | 1.505; 3.277 |
|  | IgM-MBP | 1.035 | 0.199 | 27.072 | <0.001 | 2.815 | 1.906; 4.157 |
| **BEST PREDICTION USING ALL IgA/IgG/IgM data at neuronal proteins and other** |  |  |  |  |  |  |  |
| 14 | IgG-MBP | 0.782 | 0.287 | 7.445 | 0.006 | 2.186 | 1.246; 3.833 |
|  | IgM-MBP | 1.501 | 0.284 | 27.865 | <0.001 | 4.487 | 2.570; 7.836 |
|  | IgG-SARS-CoV-2 | 0.899 | 0.229 | 15.402 | <0.001 | 2.457 | 1.568; 3.848 |
|  | IgG-HHV6 | 0.537 | 0.252 | 4.543 | 0.033 | 1.712 | 1.044; 2.805 |
|  | IgA-Activin A | 1.869 | 0.312 | 35.773 | <0.001 | 6.480 | 3.513; 11.956 |
|  | IgA to viral antigens | -2.282 | 0.340 | 44.954 | <0.001 | 0.102 | 0.052; 0.199 |

OR: Odd's ratio, 95 CI: 95% confidence intervals, Ig: Immunoglobulin
BBD: blood-brain-barrier and brain damage; MBP: myelin basic protein; MOG: myelin oligodendrocyte glycoprotein; BRA: overall index of brain reactive autoantibodies; HHV-6: Human Herpesvirus-6.

Multivariate binary regression analysis (regression #13) shows that IgG and IgM against MBP are the best predictors of Long COVID with an overall effect size of 0.303. Furthermore, another multivariate binary regression analysis which includes all IgG/IgM/IgA levels measured in our previous study (Vojdani, Almulla et al. 2023) (regression #14) indicates that a model encompassing IgG-MBP, IgM-MBP, IgG-SARS-CoV-2, IgG-HHV6 and IgA-Activin A (all positively associated), combined with IgA to viral antigens (negative association) was the most robust predictor for Long COVID disease, exhibiting a substantial effect size of 0.646. Age, sex, and research site were not significant in any of the above regression analyses.

### Intercorrelations between neuronal autoantibodies and the Long COVID phenome

**Table 3** displays that PBT has significant and positive correlations with IgA-tubulin, IgA-BBD, IgA-CP2, IgA-MBP, IgG-MBP, IgA-MOG, IgG-MOG, IgA-NFP, IgA-synapsin, and IgA-BRA. We found that IgG-MBP and IgG-MOG showed significant positive correlations with the total BDI, HAMD and HAMA and FF scores. Additionally, IgA-MBP showed significant positive correlations with BDI, HAMD, and HAMA. Moreover, there are significant and positive correlations between IgA-CP2 and BDI and HAMA. IgG-BRA showed positive and significant correlations with HAMA and FF, while IgG-synapsin displayed a significant positive correlation with the HAMA score. We also detected that the IgM-SARS-CoV-2 and IgM-HHV-6 antibodies were significantly correlated (p<0.05) with IgA, IgG and IgM directed at tubulin, IgG/IgM-BBD, IgA/IgG/IgM-CP2, IgM-MOG, IgA/IgM-NFP, IgG/IgM-synapsin, and IgG-BRA.

**Table 3.** Intercorrelation matrix between brain reactive autoimmunity and peak body temperature (PBT) and neuro-psychiatric rating scales.

| **Biomarkers** | **PBT** | **BDI** | **HAMD** | **HAMA** | **FF** |
| --- | --- | --- | --- | --- | --- |
| IgA-tubulin | 0.247<sup>*</sup> | 0.144 | 0.071 | 0.117 | 0.101 |
| IgA-BBD | 0.309<sup>**</sup> | 0.142 | 0.036 | 0.162 | -0.059 |
| IgA-cerebellar-protein-2 | 0.391<sup>**</sup> | 0.344<sup>**</sup> | 0.221 | 0.263<sup>*</sup> | 0.075 |
| IgA-MBP | 0.470<sup>**</sup> | 0.395<sup>**</sup> | 0.262<sup>*</sup> | 0.375<sup>**</sup> | 0.197 |
| IgG-MBP | 0.240<sup>*</sup> | 0.316<sup>**</sup> | 0.282<sup>*</sup> | 0.278<sup>*</sup> | 0.262<sup>*</sup> |
| IgA-MOG | 0.330<sup>**</sup> | 0.188 | 0.055 | 0.174 | -0.024 |
| IgG-MOG | 0.253<sup>*</sup> | 0.340<sup>**</sup> | 0.362<sup>**</sup> | 0.363<sup>**</sup> | 0.367<sup>**</sup> |
| IgA-NFP | 0.337<sup>**</sup> | 0.194 | 0.046 | 0.093 | -0.023 |
| IgA-synapsin | 0.336<sup>**</sup> | 0.185 | 0.046 | 0.074 | 0.030 |
| IgG-synapsin | 0.073 | 0.057 | 0.095 | 0.246<sup>*</sup> | 0.085 |
| IgG-BRA | 0.121 | 0.202 | 0.183 | 0.255<sup>*</sup> | 0.234<sup>*</sup> |
| IgA-BRA | 0.378<sup>**</sup> | 0.221 | 0.080 | 0.151 | 0.035 |

<sup>\*\*</sup>Correlation significant at $p < 0.01$ (2-tailed); <sup>\*</sup>Correlation significant at $p < 0.05$ (2-tailed).
PBT: Peak body temperature, Ig: Immunoglobulin, MOG: Myelin oligodendrocyte glycoprotein, MBP: Myelin basic protein, BBD: blood-brain-barrier and brain damage, NFP: neurofilament protein, BRA: composite of brain reactive autoantibodies

### Neuronal autoimmune responses predict the phenome of Long COVID

**Table 4** presents the results of multiple regression analyses with FF, BDI, HAMD and HAMA scores as the dependent variables and immunological markers, AOPP, CRP, HOMA2IR, age, sex, and educational level as the independent variables. Regression #1 shows that a large proportion of variance (41.7%) in FF scores could be predicted by CRP, IgG-MOG and AOPP (all positively associated). **Figure 2** shows the partial regression of the FF score on IgG-MOG. A significant part of the variance in BDI scores (37.4%) could be predicted by CRP, IgA-MBP, education and IgG-MOG (all were positively associated), as shown in regression #2. We also found in regression #3 that both CRP and IgG-MOG, which are positively associated, could significantly predict 36.6% of the variance in HAMD scores. Regression #4 reveals that IgA-MBP, CRP and education could predict a part of the variance in HAMA scores (30.4%). **Figure 3** shows the partial regression of the total HAMA score on IgA-MBP.

**Figure 2.**
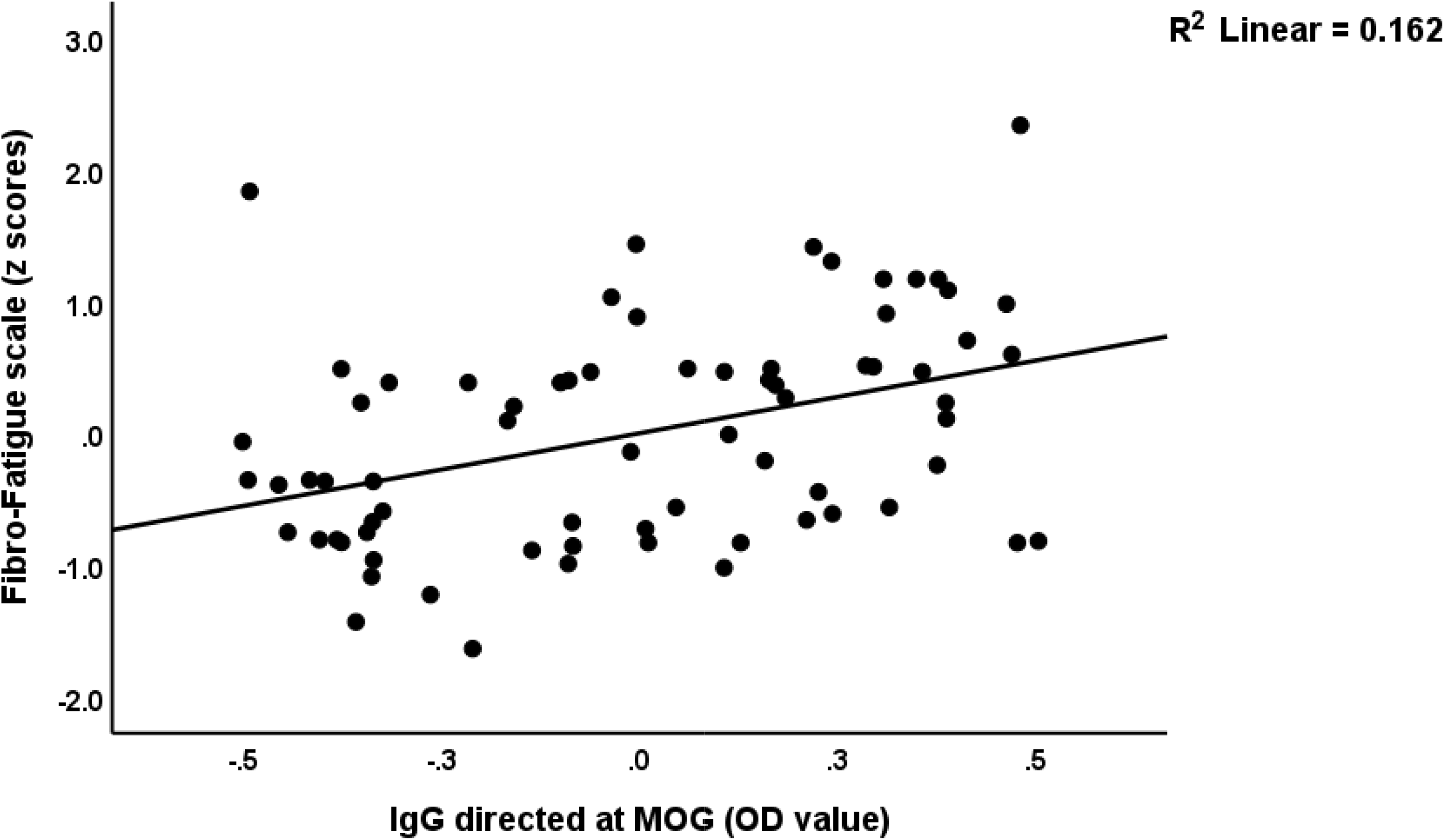
Partial regression of the Fibro-Fatigue scale score on IgG directed at myelin oligodendrocyte glycoprotein (MOG) levels.

**Figure 3.**
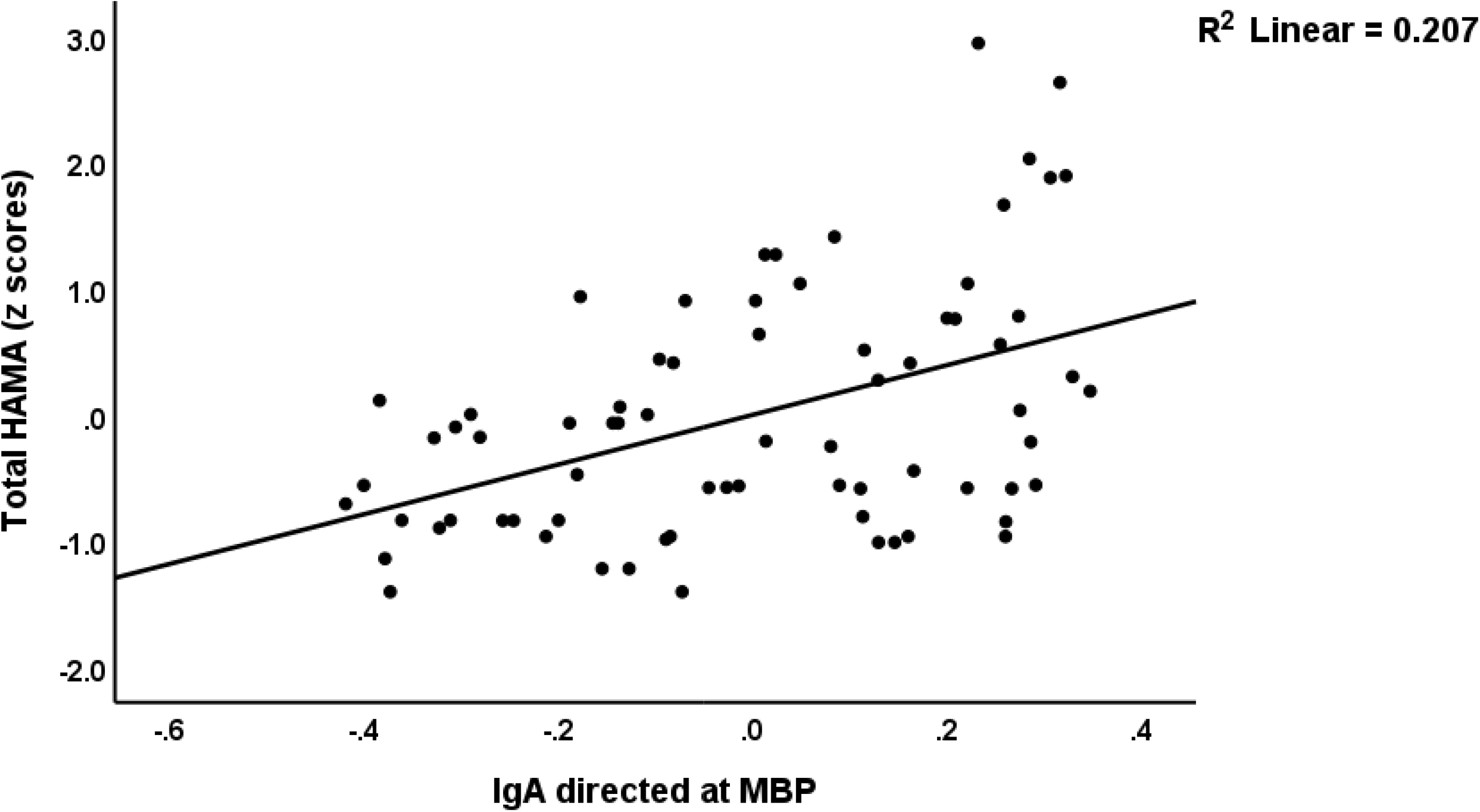
Partial regression of the total Ha directed at myelin basic protein (OD value ilton Anxiety Rating Scale (HAMA) score on IgA)

**Table 4.** Results of multiple regression analysis with the Fibro-Fatigue (FF), Beck Depression Inventory (BDI), Hamilton Depression (HAMD) and Hamilton Anxiety (HAMA) Rating Scale scores as dependent variables, and Immunoglobulins (Ig) directed at neuronal proteins, C-reactive protein (CRP), and advanced oxidative protein products (AOPP) as explanatory variables.

| Dependent Variables | Explanatory Variables | Coefficients of input variables |  |  | Model statistics |  |  |  |
| --- | --- | --- | --- | --- | --- | --- | --- | --- |
|  |  | B | t | p | R<sup>2</sup> | F | df | p |
| #1. FF_ Total | Model |  |  |  | 0.417 | 16.221 | 3/68 | <0.001 |
|  | CRP | 0.364 | 3.756 | <0.001 |  |  |  |  |
|  | IgG_MOG | 0.341 | 3.632 | <0.001 |  |  |  |  |
|  | AOPP | 0.293 | 3.051 | 0.003 |  |  |  |  |
| #2. BDI_Total | Model |  |  |  | 0.374 | 10.009 | 4/67 | <0.001 |
|  | CRP | 0.387 | 3.777 | <0.001 |  |  |  |  |
|  | IgA_MBP | 0.254 | 2.468 | 0.016 |  |  |  |  |
|  | Education | 0.218 | 2.161 | 0.034 |  |  |  |  |
|  | IgG_MOG | 0.216 | 2.092 | 0.040 |  |  |  |  |
| #3. HAMD | Model |  |  |  | 0.363 | 19.657 | 2/69 | <0.001 |
|  | CRP | 0.499 | 5.141 | <0.001 |  |  |  |  |
|  | IgG_MOG | 0.274 | 2.824 | 0.006 |  |  |  |  |
| #4. HAMA | Model |  |  |  | 0.304 | 9.896 | 3/68 | <0.001 |
|  | IgA_MBP | 0.338 | 3.268 | 0.002 |  |  |  |  |
|  | CRP | 0.363 | 3.408 | 0.001 |  |  |  |  |
|  | Education | 0.221 | 2.113 | 0.038 |  |  |  |  |

MOG: Myelin oligodendrocyte glycoprotein, MBP: Myelin basic protein

### Results of neural network analysis

We also examined the influence of the neuronal autoimmune responses, inflammatory and oxidative stress biomarkers, age, sex, and education on the CFS and affective symptoms of Long COVID using neural network analysis. **Table 5** (NN#1) utilizes the FF score as the dependent variable. In this model, both the hidden and output layers adopted the hyperbolic tangent as their activation functions, and the training was conducted with two hidden layers, each containing one unit. During the training phase, the error term was reduced, suggesting enhanced trend generalization by the neural network model. Given the consistent relative error terms across training, testing, and holdout samples, the model appears to have minimized overfitting. The cross validated precision of the model (predicted versus observed value) was 0.721. **Figure 4** illustrates the importance indicating the importances of each input variable. The model predominantly identified IgG-MBP, CRP, AOPP, IgA-synapsin, IgM-synapsin, and IgG-BRA as having the strongest predictive capacities, while variables like sex, education, and IgA-CP2 were less influential.

**Figure 4.**
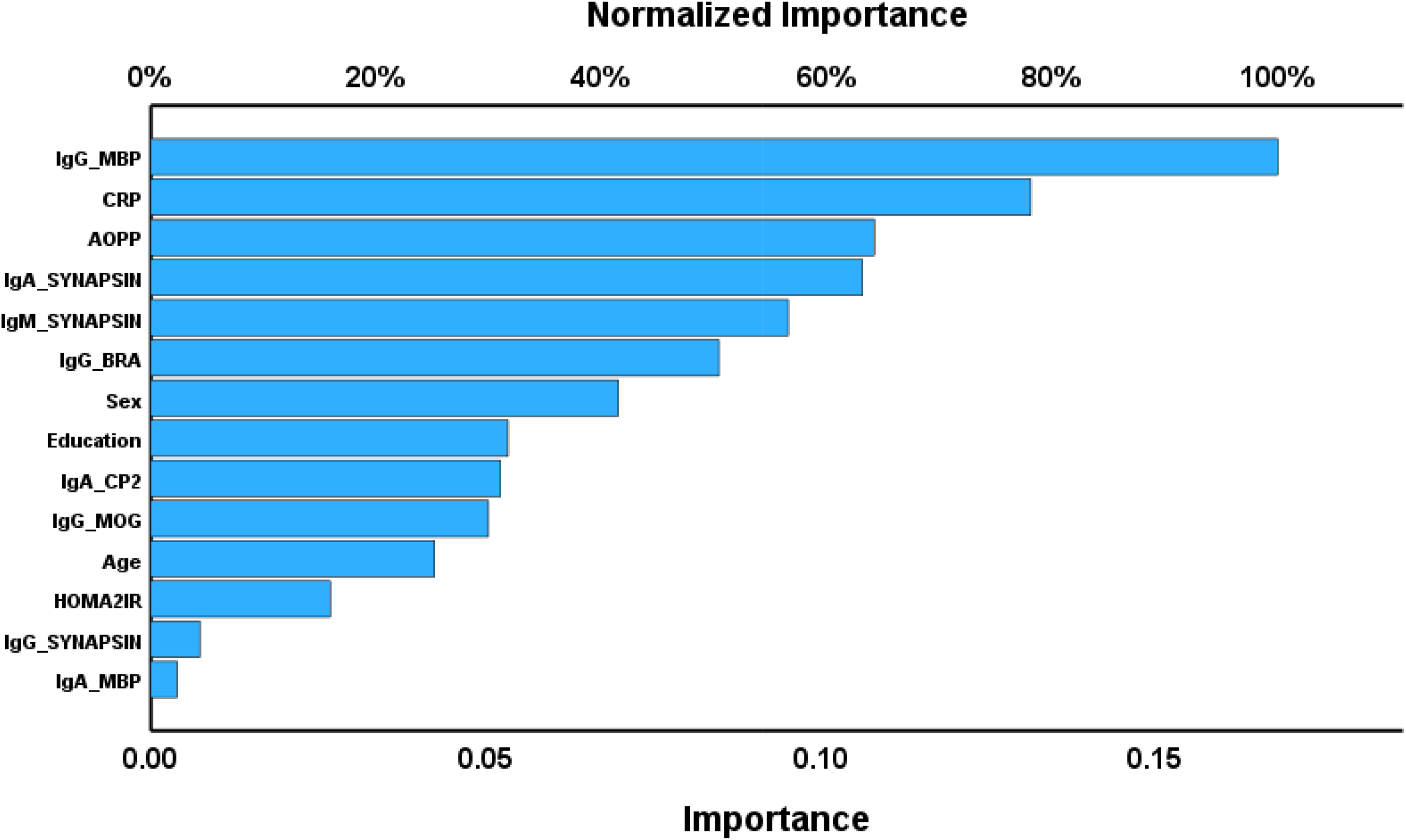
Importance chart of a neural network analysis with Fibro-Fatigue total scores as output variable mmunoglobulin. MBP: Myelin basic protein, CRP: C-reactive protein, AOPP: Advanced oxidative pro roducts, BRA: Brain-reactive autoimmunity, CP2: cerebellar-protein-2, MOG: Myelin oligodendro lycoprotein, HOMA2IR: Homeostatic model assessment insulin resistance.

**Table 5.** Results of neural networks (NN) with the chronic fatigue and affective symptoms of the Long COVID as output variables and immunological parameters as input data.

|  | **Models** | **NN#1** | **NN#2** |
| --- | --- | --- | --- |
| **Hidden layers** | Number of hidden layers | 2 | 2 |
|  | Activation function | Hyperbolic tangent | Hyperbolic tangent |
|  | Number of units in hidden layer 1 | 1 | 2 |
|  | Number of units in hidden layer 2 | 1 | 2 |
| **Output layer** | Dependent variable | Total FF score | Comp_affective |
|  | Number of units | 1 | 1 |
|  | Activation function | Hyperbolic tangent | Hyperbolic tangent |
| **Training** | Error term (sum of squares) | 2.801 | 1.221 |
|  | relative error | 0.481 | 0.340 |
| **Testing** | Sum of Squares error | 1.404 | 0.778 |
|  | relative error | 0.526 | 0.507 |
| **Holdout** | relative error | 0.821 | 0.694 |
|  | r value (predicted vs observed) | 0.721 | 0.801 |

FF: Fibro-Fatigue score; Comp\_affective: a composite extracted from depression and anxiety rating scale scores.

NN#2 shows another neural network with the Comp_affective score (z scores based on HAMD, HAMA and BDI) as dependent variable. The activation functions utilized for the hidden and output layers in this model were hyperbolic tangent. The model was trained using a configuration consisting of two hidden layers, each containing 2 units. The cross-validated precision (correlation coefficient indicating the relationship between the predicted and observed values) was 0.801. **Figure 5** is a relevance chart showing the input variables’ normalized importance. The model assigned the most predictive power to CRP, IgM-synapsin, education, IgA-MBP, IgG-MOG, IgA-synapsin, IgA-CP2, and IgG-MBP. On the other hand, IgG-BRA, sex, and age showed a less significant predictive power.

**Figure 5.**
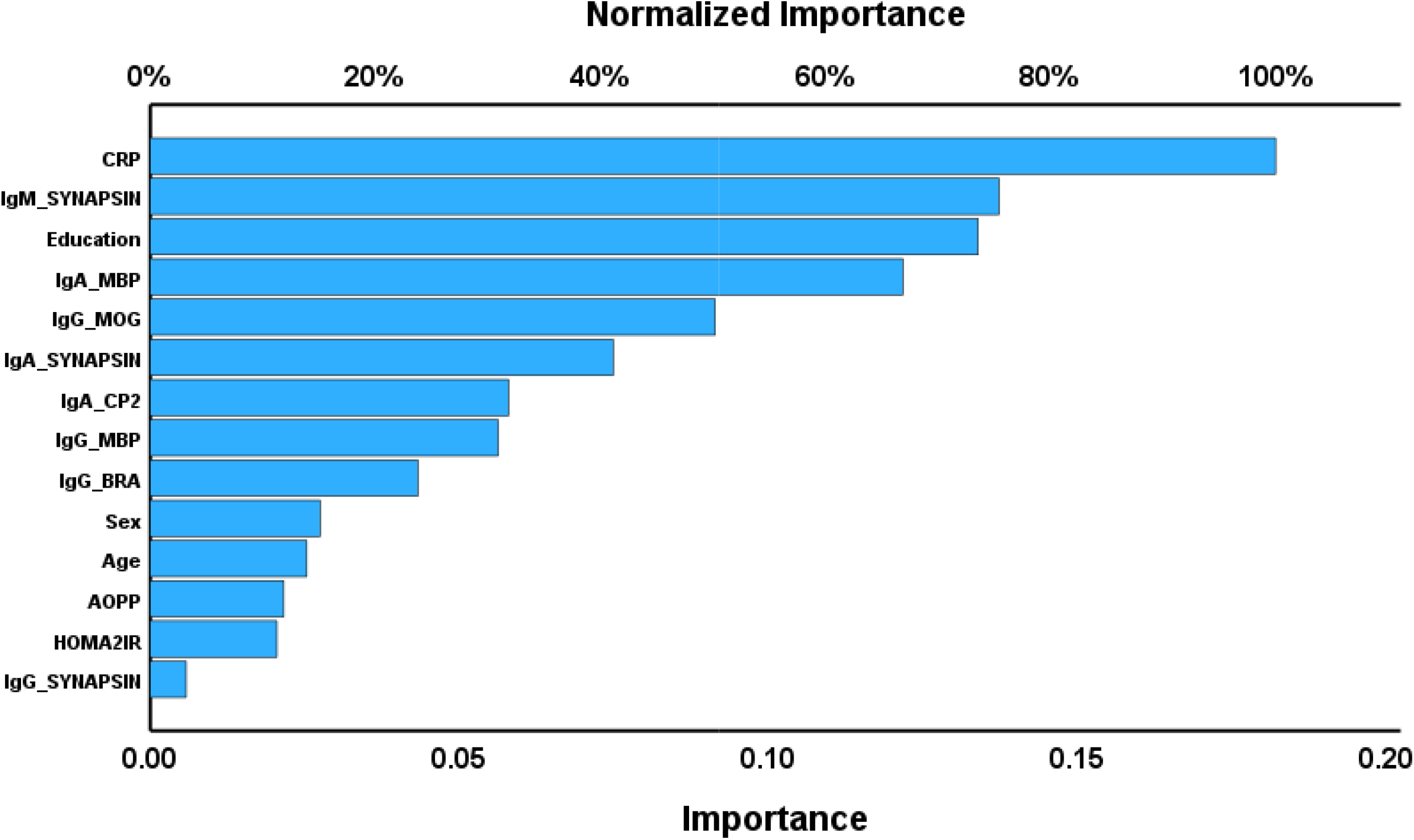
Importance chart of a neural network analysis with (Hamilton Anxiety Rating Scale + Hamil epression Rating Scale + Beck Depression Inventor scores) as output variable. CRP: C-reactive protein, M yelin basic protein, MOG: Myelin oligodendrocyte glycoprotein, CP2: Cerebellar-protein-2, BRA: Brain-reac toimmunity, AOPP: Advanced oxidative protein products, HOMA2IR: Homeostatic model assessment insulin resistance

## Discussion

### Activated immune system and increased autoimmunity in Long COVID disease

In the present study, the first significant finding is that Long COVID disease is accompanied by substantial upregulated autoimmune, inflammatory, and oxidative processes as indicated by high levels of autoimmune responses directed at diverse neuronal antigens along with increased CRP and AOPP in patients compared to healthy controls. These results extend those of our previous reports showing that Long COVID patients have significantly higher levels of autoantibodies against Activin A, as well as of upregulated immune-inflammatory and oxidative/nitrosative stress pathways (Al-Hakeim, Al-Rubaye et al. 2022, Al-Hakeim, Al-Rubaye et al. 2022, Al-Hakeim, Abed et al. 2023, Al-Hakeim, Al-Rubaye et al. 2023, Al-Hakeim, Khairi Abed et al. 2023, Almulla, Al-Hakeim et al. 2023, Vojdani, Almulla et al. 2023).

Some prior studies have reported that autoimmune reactions targeting peripheral proteins and nucleic acids may play a role in Long COVID (Lui, Lee et al. 2021, Rojas, Rodríguez et al. 2022, Son, Jamil et al. 2023, Vojdani, Almulla et al. 2023). Many patients recovering from COVID exhibit increased IgG levels directed against several self-proteins, notably IFN-α, histone, and centromere protein (Rojas, Rodríguez et al. 2022). A high percentage of Long COVID patients have latent autoimmunity and poly-autoimmunity (Rojas, Rodríguez et al. 2022). Moreover, Lui et al, detected that Long COVID patients have significantly increased antibodies against thyroid peroxidase (Lui, Lee et al. 2021). Additionally, Son et al. found that 33% of patients had developed autoreactive IgG antibodies three months after infection, and that over 40% of these are anti-ds DNA and anti-SS-B/La (Son, Jamil et al. 2023). Autoantibodies to G protein-coupled receptors (GPCRs) (such as alpha1-and alpha2-adrenoceptors, the angiotensin receptor, the nociception-like opioid receptor, and the muscarinic M2-receptor) are detected in Long COVID disease (Wallukat, Hohberger et al. 2021). The detection of GPCR autoantibodies is associated with decreased peripapillary vessel density in the eye, a known indicator of microcirculatory health (Szewczykowski, Mardin et al. 2022). The highest autoantibody levels were seen in patients treated in intensive care units, followed by those admitted to medical wards, and finally, those who recuperated at home (Don 2023).

Nevertheless, the current study refers explicitly to increased autoantibodies to neuronal antigens in association with neuro-psychiatric symptoms as clinical manifestations of Long COVID.

### Prediction of Long COVID by brain reactive autoimmune responses

The second significant finding in the current study is that the diagnosis of Long COVID disease could be externally validated by autoimmune biomarkers (IgA/IgM/IgG) directed to different CNS proteins and that addition of antibody levels directed to SARS-CoV-2 (IgG and IgM), HHV-6 (IgM and IgG), and IgA to Activin A further improve this prediction. These results indicate that the clinical diagnosis of Long COVID could be externally validated when we combine biomarkers of autoimmunity targeting CNS proteins, viral persistence, and viral reactivation. These findings confirm the hypothesis that the pathophysiology of Long COVID disease involves the persistence of SARS-CoV-2 infection, reactivation of dominant viruses, and autoimmunity (Vojdani, Vojdani et al. 2023).

Moreover, we detected that many of the autoantibody levels towards neuronal proteins were significantly predicted by the PBT obtained during the acute infection phase. Thus, increased PBT significantly predicted IgA levels directed at tubulin, CP2, MBP, MOG, NFP, and synapsin, and additionally IgG directed at MBP and MOG. Phrased differently, the severity of the inflammation during the acute phase appears to play a role in the onset of autoimmune responses to neuronal antigens which are associated with Long COVID disease. In this respect, it is worth mentioning that PBT values also predict the severity of the physio-affective phenome of Long COVID, as has been shown previously (Al-Hadrawi, Al-Rubaye et al. 2022). In addition, increased PBT and lowered SpO2 predict the activated immune-inflammatory and oxidative pathways that are observed in Long COVID (Al-Hakeim, Al-Rubaye et al. 2022).

Moreover, we found significant correlations between increased IgM levels to SARS-CoV-2, HHV-6 and HHV-6-duTPase (which are all increased in Long COVID) and diverse IgM and IgG autoantibodies directed to neuronal antigens. As such, our results show that the severity of the acute phase, SARS-CoV-2 persistence and HHV-6 reactivation all play a part in the autoimmune pathophysiology of Long COVID.

### Autoimmune antibodies to neuronal proteins in Long COVID

MBP, integral to myelin sheaths, is pivotal in myelin development and is often termed the “chief protein of the myelin sheath.”(Zhang, Sun et al. 2014). After a brain injury, MBP detaches from the plasma membrane and is released into the extracellular matrix. Here, it acts as an antigen, triggering immune-inflammatory responses (Zhang, Sun et al. 2014). Furthermore, MBP has the potential to attach to the outer layer of neuronal plasma membranes, leading to neurotoxic effects by destabilizing lipid bilayers and enhancing membrane permeability. Consequently, MBP might contribute to demyelination in post-brain injury and associated inflammatory responses and cellular death (Zhang, Sun et al. 2014). IgG antibodies targeting MBP exhibit hydrolytic, proteolytic, or abzyme actions against MBP, consequently resulting in the breakdown of the myelin sheath and triggering immune-inflammatory responses (Warren and Catz 1999, Ponomarenko, Durova et al. 2006). Elevated levels of autoantibodies against MOG have been detected in various demyelinating conditions, encompassing optic neuritis, transverse myelitis, acute disseminated encephalomyelitis, and cerebral cortical encephalitis. These conditions are now collectively acknowledged under the umbrella term MOG antibody-associated diseases (MOGAD) (Marignier, Hacohen et al. 2021). Therefore, the current results suggest that some subgroups of Long COVID patients may perhaps be considered as MOGADs.

Synapsin I autoantibodies, previously identified in individuals with neurological disorders, have been demonstrated to modulate synaptic transmission and influence neuronal synaptic density (Rocchi, Sacchetti et al. 2019). It has been reported that motor impairment, intellectual incapacity, and epilepsy are the principal clinical signs of a wide range of brain abnormalities caused by mutations in tubulin genes. These disorders manifest as aberrant neuronal migration, structure, differentiation, and axon guidance (Binarová and Tuszynski 2019).

### Autoimmune-inflammatory responses predict the phenome of Long COVID

The third significant finding of this study is that the severity of the physio-affective phenome of Long COVID disease, as defined by CFS, depression, and anxiety symptoms, could be significantly predicted by IgA/IgG/IgM responses against neuronal proteins, and increased serum CRP and AOPP levels. Increased CRP, IgA-MBP, and IgG-MOG explain a large part of the variance in the physio-affective phenome of Long COVID, 3-6 months after COVID recovery.

Previous papers have observed the presence of autoimmunity in CFS, depression, and anxiety (Maes, Mihaylova et al. 2012, Maes, Kubera et al. 2013, Maes, Ringel et al. 2013, Morris, Berk et al. 2014, Almulla, Thipakorn et al. 2023). More importantly, damage to neuronal antigens has been established in major depression (Al-Hakeim, Al-Naqeeb et al. 2023) and ME/CFS (Morris and Maes 2013), two major dimensions of Long COVID. Alterations in αβ-tubulin dimers and γ-tubulin have been implicated in brain malformations and are recognized contributors to cognitive dysfunctions (Yuba-Kubo, Kubo et al. 2005, Keays, Tian et al. 2007), which are other features of depression and ME/CFS (brain fog). Individuals with longstanding CFS have significantly higher levels of antibodies directed at microtubulin-associated protein-2 (Vernon and Reeves 2005). In MDD and schizophrenia, the observed interruption of physiological connections within the brain resulting from obligatory anomalous cytoskeletal organization was attributed to post-translational modification alterations of tubulin (Wong, Chang et al. 2013). Interestingly, neuropsychiatric systemic lupus erythematosus patients show significantly elevated antibodies against α-tubulin (Ndhlovu, Preuß et al. 2011).

In summary, the physio-affective phenome of Long COVID disease is accompanied by activation of immune-inflammatory responses, SARS-CoV-2 persistence, HHV-6 reactivation which together may lead to the formation of autoantibodies targeting neuronal proteins (Vojdani, Vojdani et al. 2023).

These findings indicate that patients with Long COVID disease are prone to increased neurotoxicity, as indicated by increased autoimmune responses to CNS antigens, probably as a consequence of infection (either persistence or reactivation), increased inflammation, and oxidative stress toxicity (Al-Hakeim, Al-Rubaye et al. 2022). As such the development of neurodegenerative disease in people with Long COVID (Elizalde-Díaz, Miranda-Narváez et al. 2022) may be explained, in part, by the increased autoantibodies directed to numerous CNS regions. In this respect, a recent meta-analysis found an association between SARS-CoV-2 infection and new onset of Alzheimer’s disease, dementia, and Parkinson’s disease in individuals post-recovery (Rahmati, Yon et al. 2023).

### Limitations

When evaluating the present findings, it’s essential to recognize specific constraints. It would be more essential to follow up the patients to examine the possible consequences of the autoimmune responses. Future studies should examine other autoantibodies to neuronal self-antigens in Long COVID and the same panel in the acute stage of COVID illness. Given that autoimmune reactions against lipid-derived neoepitopes (oxidatively modified neoepitopes) resulting from oxidative stress were recently reported in affective disorders and ME/CFS (Maes, Mihaylova et al. 2006, Almulla, Thipakorn et al. 2023), studies addressing this kind of autoimmune responses would be highly significant.

## Conclusions

The current study provides the first evidence indicating that autoimmune responses directed at neuronal proteins play a key role in Long COVID disease. Brain reactive autoantibodies directed at MBP, MOG, tubulin, CP2, and synapsin are elevated in patients with Long COVID disease indicating a neuro-autoimmune pathophysiology of this condition. The severity of the physio-affective phenome (CFS, depressive and anxiety dimensions), which represents a major dimension of Long COVID, is significantly predicted by increased IgM/IgA-synapsin, IgA/IgG-MBP, IgG-MOG, and CRP and AOPP levels. The current study is a proof of concept and mechanism study that autoimmune, inflammatory, and oxidative responses play a significant role in the pathophysiology of Long COVID disease, and in the CFS and affective symptoms (the physio-affective phenome) due to Long COVID.

## Data Availability

Upon receiving a suitable request and after the author has thoroughly utilized the data, the lead author (MM) is willing to provide access to the SPSS file associated with this study.

## Acknowledgments

The authors extend their appreciation to the contributors from all centers and hospitals mentioned in this study for their assistance in data gathering.

## Ethical approval and consent to participate

This study received approval from the Ethics Committee of the College of Medical Technology at the Islamic University of Najaf, Iraq (Document No. 34/2023). All procedures adhered to local, international, and Iraqi ethical standards. Both patients and controls provided written informed consent.

## Author’s contributions

AA and AV were responsible for blood sample collection and other patient-related tasks. Biomarker quantification in the serum was performed by AV and AFA. MM handled the statistical evaluation of the study. The manuscript was composed and refined by AFA, MM, AV, BZ and HAH, with all authors reviewing and endorsing the final version.

